# Concentrations and Estimated Daily Intake of Legacy and Emerging PFAS in the Milk of Women Residing in the Greater Cincinnati Area

**DOI:** 10.64898/2025.12.05.25341559

**Authors:** Angelico Mendy, Erin P. Hines, Aaron Dixon, Susan M. Pinney, Shannon Conrey, Hatice Cetinkaya, Mary Allen Staat, Ardythe L. Morrow

**Affiliations:** Division of Epidemiology, Department of Environmental and Public Health Sciences, University of Cincinnati College of Medicine, Cincinnati, OH, USA; Reproductive and Developmental Toxicology Branch, Public Health Integrated Toxicology Division, Center for Public Health and Environmental Assessment, Office of Research and Development, United States Environmental Protection Agency, Research Triangle Park, NC, USA; Department of Population and Quantitative Health Sciences, Case Western Reserve University School of Medicine, Cleveland, OH USA; Division of Infectious Diseases, Cincinnati Children’s Hospital, Cincinnati, OH, USA

**Keywords:** Human Milk, Lactation, Breastfeeding, Per- and Polyfluoroalkyl Substances

## Abstract

**Background:** Per- and polyfluoroalkyl substances (PFAS) have been reported in human milk. However, prior U.S. studies have not included novel PFAS alternatives of emerging concern or infants’ Estimated Daily Intake (EDI) of PFAS.

**Methods:** Human milk was collected between 2019 and 2020 at 6 weeks after delivery from 100 Cincinnati, Ohio, nursing women participants in the IMPRINT study; 29 PFAS congeners were measured using ultrahigh performance liquid chromatography-mass spectrometry. We performed descriptive exposure analyses and assessed infant’s PFAS EDI from human milk.

**Results:** All human milk samples contained PFAS. Of the 19 PFAS detected, 5 congeners were concurrently found in ≥ 50% of the samples. Legacy PFAS had the highest detection frequencies and concentrations: 97.7% for perfluorooctanesulfonic acid (PFOS) (median concentration: 14.5 ng/L) and 89.8% for perfluorooctanoic acid (PFOA) (median: 17.4 ng/L), 71.6% for perfluorohexanesulfonic acid (PFHxS) (median: 3.7 ng/L), and 70.0% for perfluorohexanoic acid (PFHxA) (median: 10.4 ng/L). An emerging PFAS, dodecafluoro-3H-4,8-dioxanonanoate (ADONA), was detected in 68.0% of samples (median: 3.5 ng/L). The PFAS with the highest EDI included PFOA (median: 8.6 ng), PFOS (median: 7.1 ng), and PFHxA (median: 5.8 ng). About 98% of samples had PFAS levels above the European Food Safety Authority (EFSA) tolerable weekly intake of 4.4 ng/kg body weight/week for the sum of PFOA, PFOS, PFHxS and perfluorononanoate (PFNA).

**Conclusions:** Human milk from women in Cincinnati, Ohio, contained both legacy and emerging PFAS and infants’ PFAS consumption through breastfeeding exceeded EFSA tolerable weekly intakes.

## Introduction

Per- and polyfluoroalkyl substances (PFAS) are fluorinated aliphatic compounds with hydrophobic and oleophobic properties used in a wide variety of commercial and consumer products (1). Drinking water containing PFAS has been a significant exposure source in the U.S., especially the Mid-Ohio Valley, the site of this study cohort (2–4). Legacy PFAS or long-chain PFAS (fluorinated carbons ≥ 8 for perfluoroalkyl carboxylates [PFCA] or ≥ 6 for perfluoroalkyl sulfonate [PFSA]) that were voluntarily phased out of production in several developed nations, include perfluorooctane sulfonic acid (PFOS) and perfluorooctanoic acid (PFOA) (1, 5, 6). These can bioaccumulate and persist in humans and wildlife as well as in the environment for decades and have been associated with carcinogenic, immunotoxic, cardiovascular, respiratory, metabolic, renal, and reproductive disorders in humans (1, 5, 6).

PFOS and PFOA are known immunotoxicants associated with reduced antibody responses to vaccines and innate immunity disruption through natural killer cells, B-lymphocytes and neutrophil cell count decreases (6, 7). As a result of these deleterious effects and in response to concerns about potential impacts to human health, 3M began to voluntarily phase out PFOA and PFOS by 2002, and 2006, respectively as well as similar long-chain PFAS. Likewise, the U.S. Environmental Protection Agency launched the PFOA Stewardship Program in 2006, which focused on reducing, then eliminating, PFOA from facility emissions and product content (8). Since that time, PFOA and PFOS have been measured at lower concentrations in US populations, while other PFAS chemistries have been on the rise in Western countries, where short-chain PFAS (fluorinated carbons ≤ 7 for PFCA or ≤ 5 for PFSA) have been introduced as alternatives (9, 10).

Human milk is a major source of PFAS exposure in nursing infants and may contribute up to 94% of PFAS exposure in the first six months of a child’s life (11). The American Academy of Pediatrics and the World Health Organization recommend exclusively breastfeeding children until the age of 6 months and continuing breastfeeding with the introduction of complementary foods until at least 2 years (12). Alternatively, PFAS in drinking water can become part of infant exposure through water’s use in reconstituting infant formula and is a source of maternal exposure to PFAS (12). Since young children have higher daily PFAS intakes than adults alongside immature detoxification systems and organs, prolonged early life exposure to PFAS may have unique health consequences (13, 14). Although the U.S. does not have regulatory guidance for levels of PFAS in human milk, the European Food Safety Authority (EFSA) defined critical human milk PFAS levels as 60 ng/L for PFOA and perfluorononanoate (PFNA), 73 ng/L for perfluorohexanesulfonate (PFHxS) and PFOS, and 133 ng/L for the sum of the four PFAS in 2020 (15). Due to PFAS’ ability to bioaccumulate, EFSA has also established a tolerable weekly intake (TWI) of 4.4 ng/kg body weight per week for the total of the four PFAS (PFOA, PFOS, PFFNA and PFHxS) as a threshold for potential adverse health effects (15).

Nevertheless, research on PFAS concentrations in human milk in the U.S. remains limited. (10, 16, 17). Most of the few studies conducted in the U.S. have focused on long-chain legacy PFAS, only one has included short chain PFAS and none have reported human milk content of per- and polyfluoroether carboxylates (PFECA), a novel class of legacy PFAS alternatives with emerging environmental and health concerns (10). Notable PFECA are perfluoro-2-propoxypropanoic acid (HFPO-DA or GenX) and 4,8-dioxa-3H-perfluorononanoate (ADONA); negative health effects for these PFECA molecules have been reported from *in-vitro* studies and in zebrafish and rodents (18). To address current gaps in knowledge, we aimed to examine the distribution and sociodemographic predictors of legacy and emerging PFAS in human milk among women from the greater Cincinnati, Ohio area.

## Methods

### Study Participants

One hundred breastfeeding women recruited at Cincinnati Children’s Hospital Medical Center as participants in the IMPRINT cohort study were included in this analysis. IMPRINT is a NIAID-funded pregnancy and birth cohort that used a standardized protocol for human milk collection in breastfeeding mothers at 2- and 6-weeks post-partum and annually until breastfeeding cessation. Mother-child pairs were eligible for inclusion at baseline if the woman was ≥18 years old, had a live born infant of ≥ 34 weeks, planned to deliver at a local hospital, lived within designated zip codes with no plans to move within 4 years, and had a cell phone for text messaging. Mother-child pairs were excluded from the study if they had fetal or infant death before maternal discharge from hospital, had multiples, had an identified HIV infection, congenital disorders, heroin, cocaine or methamphetamine use during pregnancy, or enrolled in a vaccine trial.

### Measurement of PFAS in Human Milk

PFAS were measured in human milk collected between 2019 and 2020 at 6 weeks after delivery using ultrahigh performance liquid chromatography-tandem mass spectrometry (UPLC-MS/MS) by SGS AXYS Analytical Services, British Columbia, Canada, under the SGS AXYS Method MLA-110 based on EPA Method 1633 (19). In brief, study samples (range of 1.2 to 2mL) were first spiked with isotopically labeled surrogate standards. Samples were extracted with methanolic potassium hydroxide solution, with acetonitrile, and finally with methanolic potassium hydroxide solution, collecting the supernatants. The supernatants were combined, treated with ultra-pure carbon powder and evaporated to remove methanol. The resulting solution was diluted with water and cleaned up by solid phase extraction on a weak anion exchange sorbent. The eluate was spiked with recovery standards and analyzed by UPLC-MS/MS. PFAS quantification was done using the isotope dilution and internal standard method, by comparing the area of the primary transition to that of the (15) C-labelled or deuterium labeled standard, correcting for response factors. Sample-specific detection limits (SDL) were determined by converting the area equivalent to three times the estimated chromatographic noise height to a concentration in the same manner that target peak responses were converted to final concentrations. We performed blank corrections for compounds that had background contribution in the blank samples, and we determined the detection limit as 3 times the standard deviation of the mean blank concentrations. Samples with PFAS concentrations below detection were given the value SDL/√2. Due to the methods development nature of this study, not all PFAS listed in Method 1633 were measured.

PFAS were categorized as PFCA, PFSA, fluorotelomer sulfonates (FTS), PFECA, and perfluorooctane sulfonamides (PFOSA). PFCA included perfluoropentanoate (PFPeA), perfluorohexanoate (PFHxA), perfluoroheptanoate (PFHpA), PFOA, PFNA, perfluorodecanoate (PFDA), perfluoroundecanoate (PFUnA), perfluorotridecanoate (PFTrDA), and perfluorotetradecanoate (PFTeDA). PFSA comprised perfluorobutanesulfonate (PFBS), perfluoropentanesulfonate (PFPeS), PFHxS, perfluoroheptanesulfonate (PFHpS), PFOS, perfluorononanesulfonate (PFNS), and perfluorodecanesulfonate (PFDS). FTS included 4:2, 6:2, and 8:2 FTS. PFECA included HFPO-DA, ADONA, and perfluoro-3,6-dioxaheptanoate (NFDHA). PFOSA were perfluorooctanesulfonamide (PFOSA), 2-N-methylperfluoro-1-octanesulfonamidoacetate (MeFOSAA), and 2-N-ethylperfluoro-1-octanesulfonamidoacetate (EtFOSAA) (20). We also classified analytes as long- (PFOA, PFNA, PFDA, PFUnA, PFHxS and PFOS) and short-chain PFAS (PFPeA, PFHxA, PFHpA and PFBS) (**Figure 1**) (21).

**Figure 1.**
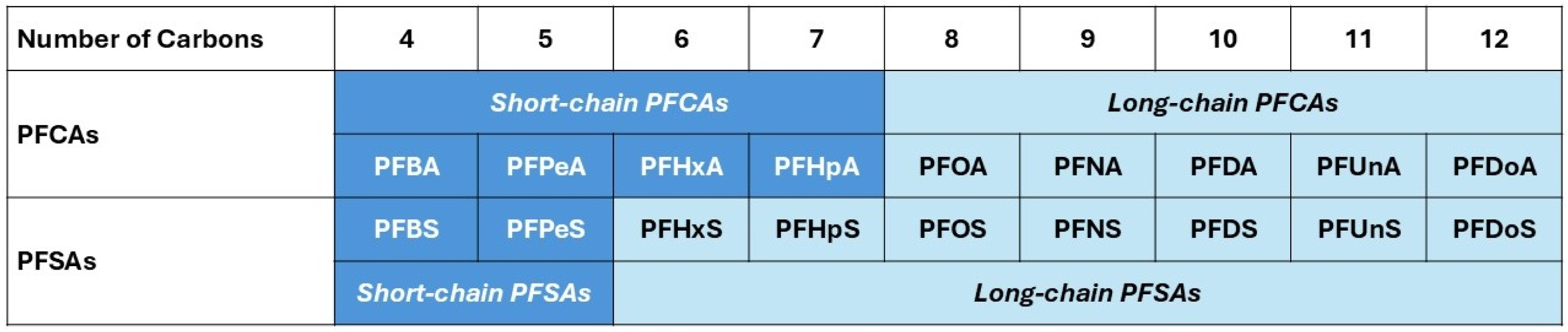
Long-chain and short-chain PFCAs and PFSAs

### Covariates

Questionnaires were administered at enrollment (3^rd^ trimester of pregnancy) to collect data on age, race, household income, educational attainment, insurance type, and parity. Women were asked about height and pre-pregnancy weight, which were used to compute body mass index (BMI). The date of sample collection was recorded to analyze the year and season of the sample collections: Fall (September to November 2019), Winter (December 2019 to February 2020), and Spring (March to May 2020).

### Statistical Analysis

We performed descriptive analyses to report the detection frequency and distribution of the 29 PFAS analyzed in the study. We reported PFAS median concentrations along with their corresponding interquartile range (IQR) overall and by participant characteristics and compared the difference in PFAS levels using the Wilcoxon Rank test. Given the skewed distribution of PFAS human milk concentrations, we applied log_10_-transformation to improve distribution normality. We generated bar graphs depicting the detection frequency of the PFAS detected in human milk and boxplots showing the concentration and distribution of PFAS detected in ≥50% of samples. The intercorrelation between the PFAS was explored using Spearman correlations, and their coefficients were interpreted as weak (ρ < 0.40), moderate (0.40 ≤ ρ < 0.70), and strong (ρ ≥ 0.70). PFAS EDI was calculated as the product of PFAS concentration and the daily volume of human milk intake infants estimated using a meta-analytic method that combined 167 studies (22). We tested whether sociodemographic characteristics (age, race, household income, educational attainment, and insurance type), number of live births, BMI, season or year of sampling, and state of residence (Ohio or Kentucky) were predictors of log_10_-transformed PFAS concentrations mutually adjusting for these variables. The linear regression diagnostic tests were performed by plotting residuals, which were normally distributed, showing that model assumptions were reasonably met. The analyses were done in RStudio (R version 4.3.2, The R Foundation, NY) and SAS (version 9.4, SAS Institute, Cary, NC) and *P* < 0.05 was considered significant.

## Results

### Characteristics of Study Participants

A complete description of study participants is presented in **Table 1**. Briefly, among the 100 study participants, the mean (standard deviation) age was 32.9 (±4.8) years; 83.0% were White and 10.0% Black. About 56.0% had a household income ≥ $75,000; 43.0% had a graduate degree and 56% were overweight or obese. Most milk samples were collected in 2020 (n=91.0) and from women residing in Southwest Ohio (85.0%). About 42% of infants were breastfed with an average age of 7.3 weeks and had a mean birthweight of 3.4 kgs.

**Table 1:** Characteristics of Study Participants, IMPRINT (N = 100)

| Characteristics | Estimate |
| --- | --- |
| Age (years), mean (SD) | 32.9 (4.8) |
| Race, % |  |
| White | 83.0 |
| Black | 10.0 |
| Other | 7.0 |
| Household income, % |  |
| <\$75,000 | 26.0 |
| ≥ \$75,000 | 56.0 |
| Missing | 18.0 |
| Education, % |  |
| No graduate degree | 57.0 |
| Graduate degree | 43.0 |
| BMI |  |
| Normal | 44.0 |
| Overweight | 29.0 |
| Obese | 27.0 |
| Insurance type, % |  |
| Public | 9.0 |
| Private | 88.0 |
| Both | 3.0 |
| Sampling Season, % |  |
| Fall | 53.0 |
| Winter | 38.0 |
| Spring | 9.0 |
| Sampling Year, % |  |
| 2019 | 9.0 |
| 2020 | 91.0 |
| State of Residence, % |  |
| Southwest Ohio | 85.0 |
| Northern Kentucky | 15.0 |
| Non-exclusive breastfeeding, % | 42.0 |
| Postpartum weeks, mean (SD) | 7.3 (0.6) |
| Birthweight (kgs), mean (SD) | 3.4 (0.5) |
| Gestational age (weeks), mean (SD) | 39.0 (1.2) |

**Abbreviations:** SD, standard deviation; BMI, body mass index

### Human Milk PFAS Detection and Levels

Five of the 19 detected PFAS had a detection frequency ≥50.0% (PFOS [97.9%], PFOA [89.8%], PFHxS [71.6%], PFHxA [70.0%], and ADONA [68.0%]) (**Table 2**). The frequency of detection of these five PFAS is depicted in decreasing order in **Figure 2**. Median total concentration of these five PFAS in human milk was 61.1 ng/L (IQR: 38.8 to 86.4) and ranged from 8.7 to 321.8 ng/L. As shown in **Figure 3** and **Table 1**, PFOA was most abundant (median: 17.4 ng/L [IQR: 11.4, 24.9]), followed by PFOS (median: 14.6 ng/L [IQR: 8.2, 21.1]), PFHxA (median: 10.4 ng/L [IQR: < LOD, 20.8]), PFHxS (median: 3.7 ng/L [IQR: < LOD, 7.0]), and ADONA (median: 3.5 ng/L [IQR: < LOD, 5.5]).

**Figure 2.**
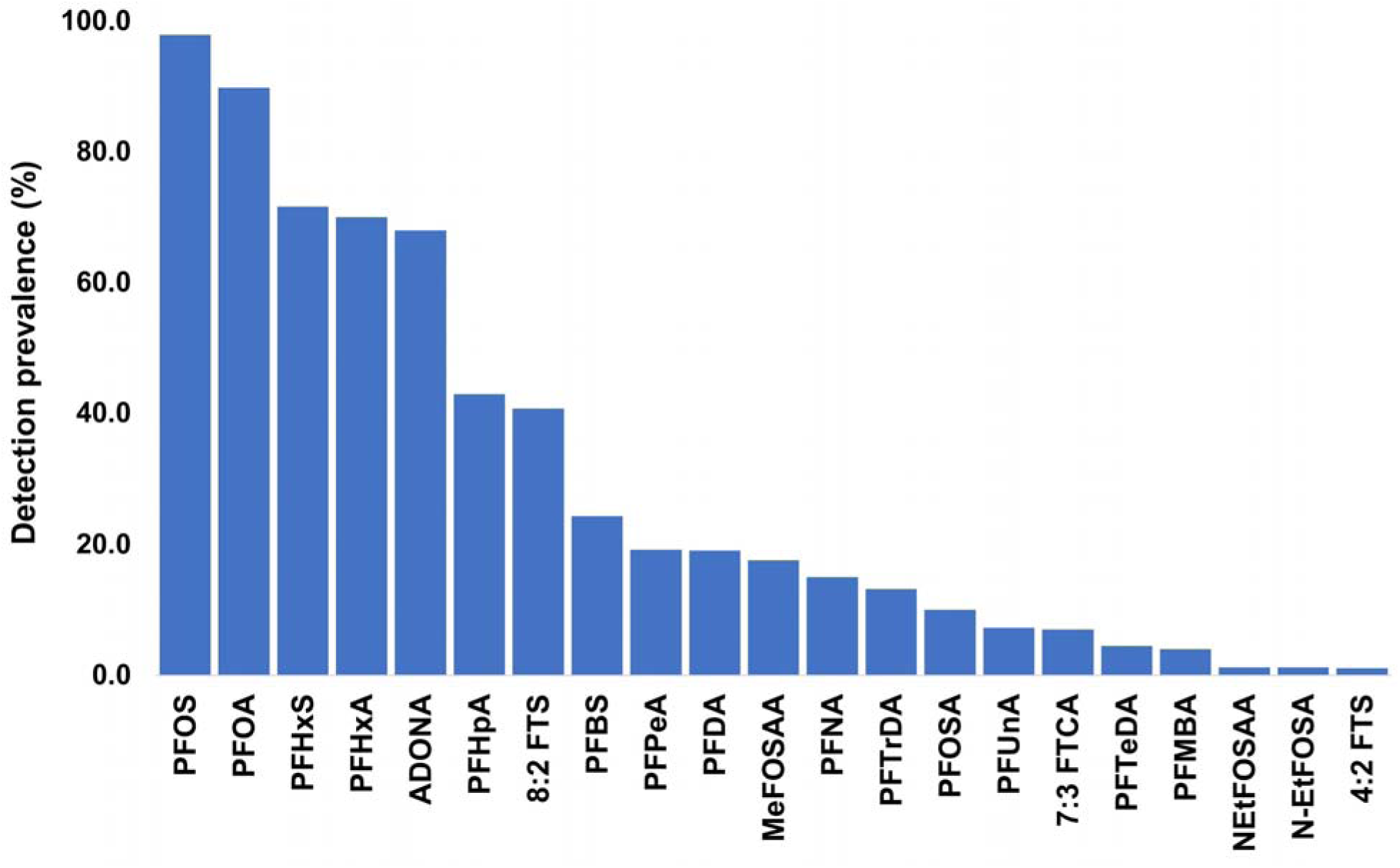
Detection prevalence of PFAS detected in human milk samples in decreasing order

**Figure 3.**
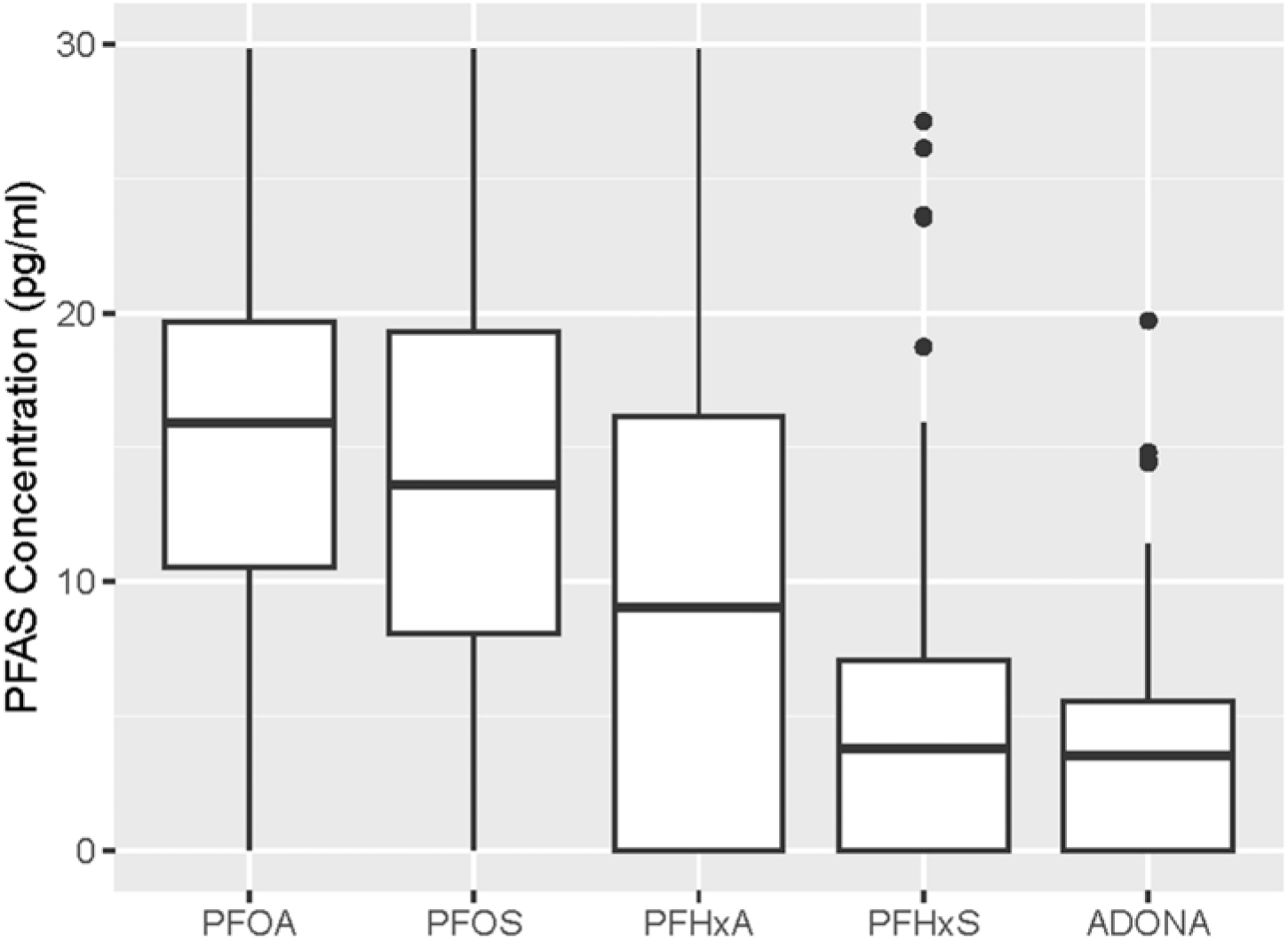
Boxplot of human milk concentrations of PFAS detected in ≥ 50% of samples showing median concentration and interquartile ranges

**Table 2:**
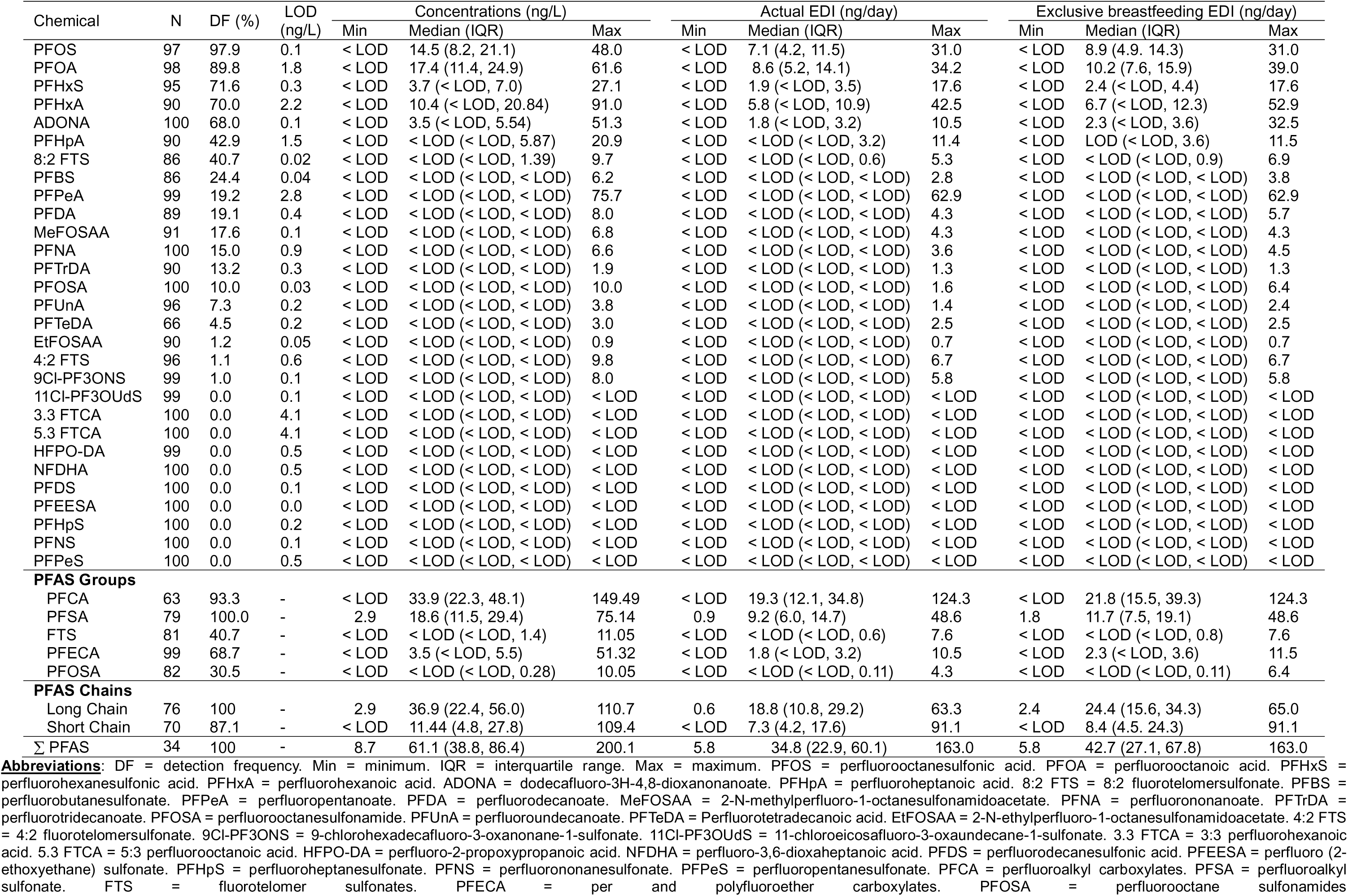
Detection frequency, concentrations (ng/L), and estimated daily intake (ng/day) of PFAS measured in IMPRINT (N = 100)

Measured concentrations of long-chain PFAS (36.9 ng/L [IQR: 22.4, 56.0]) were three times those of short-chain PFAS (11.4 ng/L [IQR: 4.8, 27.8]). The PFAS type with the highest concentration was PFCA (median: 33.9 ng/L [IQR: 22.3, 48.1]), followed by PFSA (median: 18.6 ng/L [IQR: 11.5, 29.4]), and PFECA (median: 3.5 ng/L [IQR: < LOD, 5.5]); FTS and PFOSA were detected in <50% of the samples (**Table 2**).

Only 1 out of 98 samples had a PFOA level above the EFSA critical levels of PFAS in breast milk limit of 60 ng/L, and all samples had levels below the EFSA critical levels of PFAS in breast milk limits of 60 ng/L for PFNA, 73 ng/L for PFHxS as well as PFOS, and 133 ng/L for the sum of PFOA, PFNA, PFHxS and PFOS. Even though EFSA critical levels were not exceeded, most samples (>98%) exceeded the EFSA TWI (see EDI section below).

### Human Milk PFAS Co-exposure and Intercorrelation

Half of the study participants had 5 PFAS or more concurrently detected in their human milk samples. There were moderate correlations of PFOS with PFOA and PFHxS, PFOA with PFHxS, PFHxA with ADONA, PFHpA with PFBS, PFDA and PFTrDA, PFPeA with PFTrDA, and PFNA with PFTrDA. The intercorrelations between all other PFAS were weak (**Figure 4**).

**Figure 4.**
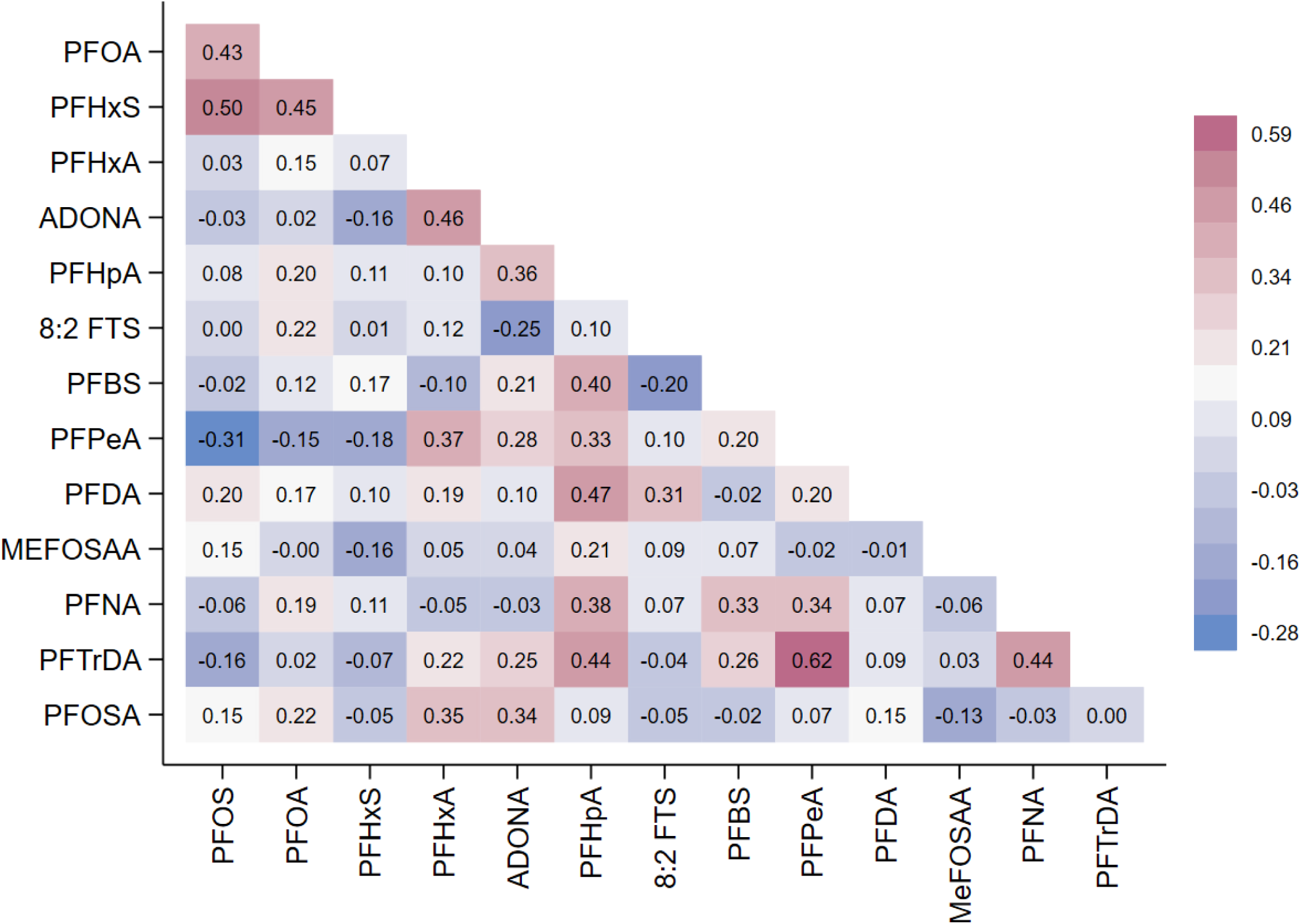
Intercorrelation of human milk PFAS detected in ≥ 50% of samples using Spearman correlation

### Estimated Daily Intake (EDI) of Human Milk PFAS

PFOA had the highest EDI (median: 8.6 ng [IQR: 5.2, 14.1]) followed by PFOS (median: 7.1 ng [IQR: 4.2, 11.5]), PFHxA (median: 5.8 ng [IQR: < LOD, 10.9]), PFHxS (median: 1.9 ng [IQR: < LOD, 3.5]), and ADONA (median: 1.8 ng [IQR: < LOD, 3.2]). The long chain PFAS EDI (median: 18.8 ng [IQR: 10.8, 29.2]) was more than twice that of short chain PFAS (median: 7.3 ng [IQR: 4.2, 17.6]). Among the PFAS groups, median EDI was 19.3 ng (IQR: 12.1, 34.8) for PFCA, 9.2 ng (IQR: 6.0, 14.7) for PFSA, and 1.8 ng (IQR: < LOD, 3.2) for PFECA, but was below detection for FTS and PFOSA. ∑PFAS median EDI was 34.8 ng (IQR: 22.9, 60.1) (**Table 2**). About 98% of the human milk samples had the sum of PFOA, PFNA, PFHxS, and PFOS ≥ EFSA TWI. Table 2 also details EDIs if public health recommendations were followed for exclusive breastfeeding at 6 months and 99% of samples were above EFSA TWI.

### Human Milk PFAS Predictors

After mutual adjustment for covariates, lower parity was associated with higher human milk PFOS (β: 0.07 [95% CI: 0.00, 0.14]), PFOA (β: 0.10 [95% CI: 0.01, 0.19]), and PFHxS (β: 0.11 [95% CI: 0.01, 0.21]). Graduate education was associated with higher PFHxS (β: 0.23 [95% CI: 0.02, 0.43]), and private insurance with higher ADONA (β: 0.36 [95% CI: 0.04, 0.67]). Compared to human milk sampled in Spring, those collected in Winter had higher PFOS (β: 0.77 [95% CI: 0.40, 1.15]), while those collected in Fall had higher PFOA (β: 0.60 [95% CI: 0.21, 0.99]), PFHxA (β: 0.67 [95% CI: 0.13, 1.21]), and ADONA (β: 0.51 [95% CI: 0.28, 0.74]). Human milk sampled in 2020 versus 2019 had higher PFOS (β: 0.18 [95% CI: 0.01, 0.35]) and PFOA (β: 0.29 [95% CI: 0.02, 0.57]). Non-exclusive breastfeeding predicted higher PFOS (β: 0.28 [95% CI: 0.12, 0.44]) and PFOA (β: 0.19 [95% CI: 0.02, 0.37]); fewer postpartum weeks predicted higher PFOA (β: 0.18 [95% CI: 0.02, 0.33]) and higher birthweight predicted higher PFOS (β: 0.25 [95% CI: 0.07, 0.43]) and PFOA (β: 0.33 [95% CI: 0.16, 0.50]) (**Tables 3**).

**Table 3:** Linear regression coefficient (β) and 95% confidence intervals for predictors of human milk PFAS, IMPRINT (N = 100)

| Characteristics | PFOS | PFOA | PFHxS | PFHxA | ADONA |
| --- | --- | --- | --- | --- | --- |
| Age (per 10 years) | 0.00 (-0.19, 0.19) | 0.10 (-0.12, 0.31) | -0.19 (-0.45, 0.07) | 0.26 (-0.17, 0.68) | -0.05 (-0.34, 0.24) |
| Race |  |  |  |  |  |
| White | Reference | Reference | Reference | Reference | Reference |
| Black | -0.18 (-0.42, 0.06) | -0.07 (-0.37, 0.23) | -0.24 (-0.67, 0.19) | 0.23 (-0.35, 0.81) | 0.06 (-0.30, 0.42) |
| Other | -0.07 (-0.22, 0.07) | -0.24 (-0.52, 0.04) | -0.07 (-0.33, 0.20) | 0.05 (-0.45, 0.56) | -0.10 (0.37, 0.17) |
| Household income |  |  |  |  |  |
| <\$75,000 | Reference | Reference | Reference | Reference | Reference |
| ≥ \$75,000 | -0.15 (-0.33, 0.04) | 0.15 (-0.10, 0.40) | 0.02 (-0.31, 0.35) | 0.02 (-0.54, 0.58) | -0.21 (-0.53, 0.11) |
| Lower parity | **0.07 (0.00, 0.14)\*** | **0.10 (0.01, 0.19) \*** | **0.11 (-0.21, 0.01) \*** | -0.10 (-0.26, 0.06) | 0.03 (-0.07, 0.14) |
| Graduate degree | 0.09 (-0.06, 0.23) | 0.08 (-0.07, 0.24) | **0.23 (0.02, 0.43) \*** | -0.17 (-0.56, 0.22) | -0.05 (-0.29, 0.19) |
| BMI (per 10) | -0.04 (-0.13, 0.05) | -0.13 (-0.27, 0.01) | 0.05 (-0.07, 0.17) | 0.13 (-0.10, 0.35) | 0.09 (-0.03, 0.21) |
| Private health insurance | 0.01 (-0.19, 0.21) | 0.15 (-0.20, 0.50) | 0.15 (-0.21, 0.51) | 0.21 (-0.36, 0.78) | **0.36 (0.04, 0.67) \*** |
| Sampling Season |  |  |  |  |  |
| Spring | Reference | Reference | Reference | Reference | Reference |
| Winter | 0.23 (-0.13, 0.58) | **0.77 (0.40, 1.15) \*\*\*\*** | 0.15 (-0.08, 0.38) | 0.31 (-0.29, 0.89) | 0.15 (-0.13, 0.43) |
| Fall | 0.11 (-0.26, 0.47) | **0.60 (0.21, 0.99) \*\*** | 0.09 (-0.15, 0.32) | **0.67 (0.13, 1.21) \*** | **0.51 (0.28, 0.74) \*\*\*\*** |
| Sampling year |  |  |  |  |  |
| 2019 | Reference | Reference | Reference | Reference | Reference |
| 2020 | **0.18 (0.01, 0.35) \*** | **0.29 (0.02, 0.57) \*** | 0.08 (-0.28, 0.43) | 0.30 (-0.23, 0.82) | 0.15 (-0.15, 0.46) |
| Residence |  |  |  |  |  |
| Southwest Ohio | Reference | Reference | Reference | Reference | Reference |
| Northern Kentucky | -0.05 (-0.24, 0.14) | -0.18 (-0.46, 0.10) | 0.10 (-0.17, 0.36) | -0.07 (-0.59, 0.43) | 0.19 (-0.09, 0.46) |
| Non-exclusive breastfeeding | **0.28 (0.12, 0.44) \*\*\*** | **0.19 (0.02, 0.37) \*** | -0.10 (-0.31, 0.12) | 0.02 (-0.33, 0.38) | 0.00 (-0.24, 0.25) |
| Fewer postpartum weeks | 0.02 (-0.11, 0.14) | **0.18 (0.02, 0.33) \*** | 0.04 (-0.13, 0.20) | -0.16 (-0.41, 0.10) | -0.16 (-0.34, 0.03) |
| Birthweight | **0.25 (0.07, 0.43) \*\*** | **0.33 (0.16, 0.50) \*\*\*** | 0.12 (-0.12, 0.36) | -0.26 (-0.66, 0.14) | 0.00 (-0.24, 0.25) |
| Gestational age | -0.01 (-0.03, 0.01) | -0.01 (-0.04, 0.01) | 0.00 (-0.03, 0.03) | 0.02 (-0.04, 0.07) | -0.01 (-0.03, 0.02) |

\* $P < 0.05$ \*\* $P < 0.01$ \*\*\* $P < 0.001$ \*\*\*\* $P < 0.001$
Abbreviations: BMI = body mass index. PFOS = perfluorooctanesulfonic acid. PFOA = perfluorooctanoate. PFHxS = perfluorohexanesulfonate. PFHxA = perfluorohexanoic acid. ADONA = dodecafluoro-3H-4,8-dioxanonoic acid.

Comparisons of recent human milk PFAS study findings to our present study results are presented in **Table 4**. For each cohort represented in Table 4, multiple PFAS congeners were measured but some had no data above the limit of detection. In studies with LODs capable of measuring milk PFAS concentrations (ng/L range), comparisons across cohorts showed similar concentrations (less than an order of magnitude difference between cohorts). The cohorts represent multiple different countries and span over 20 years of time.

**Table 4:**
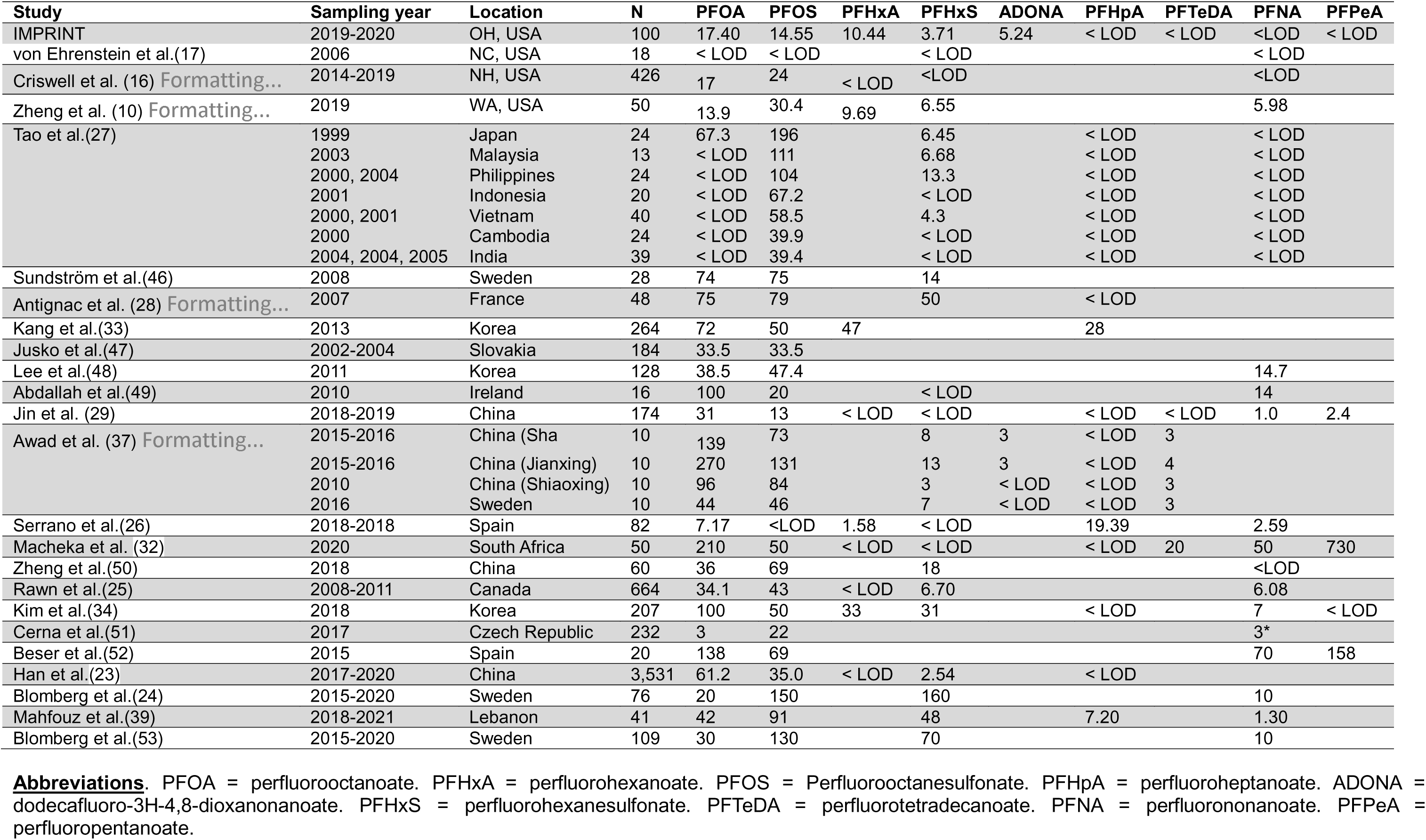
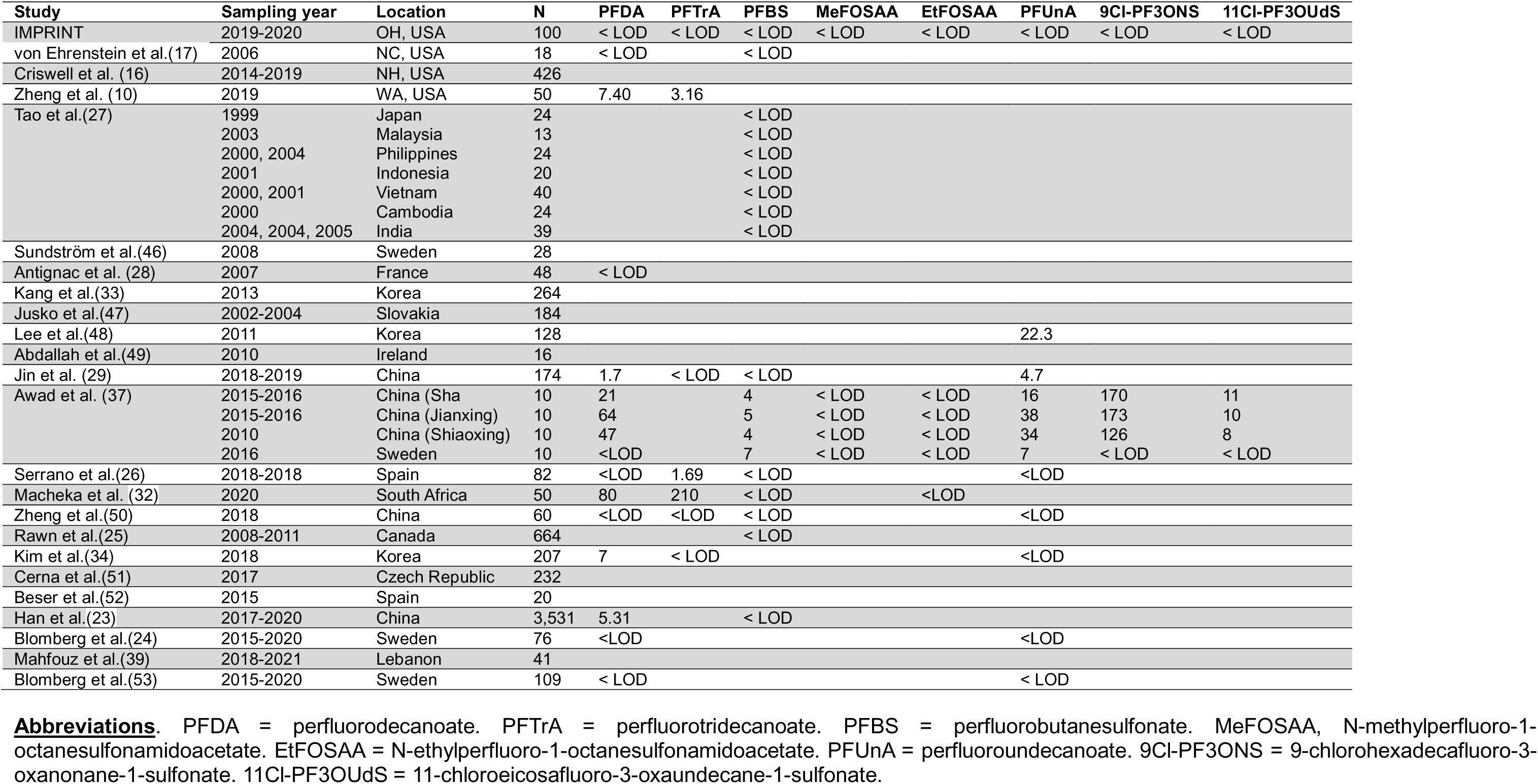
Median levels of human milk PFAS (ng/L) reported in previous studies compared to IMPRINT (N = 100)

## Discussion

All human milk sampled from our study participants contained PFAS and 5 or more PFAS congeners were concurrently found in at least half of the samples. Of the 29 PFAS measured, 19 were detected in at least one sample. The legacy PFAS, PFOS and PFOA, were the most frequently detected and had the highest concentration, while the newer PFAS such as PFHxS, PFHxA, and ADONA were widely detected in human milk samples. Infants’ intake of PFAS from human milk exceeded the EFSA TWI in 98% of participants. Depending on the congeners, predictors of higher human PFAS in milk included high socioeconomic status for PFHxS and ADONA; fewer children for PFOS, PFOA and PFHxS; Winter or Fall versus Spring season of sampling for PFOA, PFHxA and ADONA; year of sampling (2020 versus 2019); non-exclusive breastfeeding and high birthweight for PFOS and PFOA; and fewer postpartum weeks for PFOA.

### Human Milk PFAS Detection

Few U.S. studies have analyzed PFAS in human milk. Zheng et al. analyzed 50 samples collected in 2019 from Seattle, Washington (WA), women for 39 PFAS (9 short-chain and 30 long-chain) and found 16 PFAS with measurable levels above method detection limits (10). Their method detection limits (MDLs) of three times the standard deviation of the target analytes detected in blanks ranged from 0.10 ng/L (PFPeS and 8:2 FTS) to 403 ng/L (PFHxDA) (10). The study also reported that 12 out of the 16 detected PFAS were detected in more than half of their participants (10). The largest U.S. investigation included 426 samples collected between 2014 and 2019 in women from Concord and Lebanon into the New Hampshire Birth Cohort Study (NHBCS) (16). The milk samples were analyzed for 10 legacy PFAS, and the PFAS detection frequencies at LOD = 10 ng/L were 92% for PFOS and 83.6% for PFOA (16). The remaining PFAS (PFHxS, PFNA, PFDA, PFUnDA, PFBS, PFHpA, PFHpS, and PFHxA) were detected in < 8% of the samples and excluded from further analysis (16). In China, the third National Human Milk Survey from 2017 to 2020 tested for 30 PFAS (28 legacy and 2 PFESA) in 3,531 samples from 100 sites in 24 provinces (23). It found 19 PFAS detected, with PFOA and PFOS being present in all samples at LODs ranging from 1 ng/L (for PFDA, PFUdA, PFBS, PFPeS, PFHxS, and PFHpS) to 12 ng/L (for PFOA) (23). In Sweden, the Ronneby Mother-Child Cohort included 76 samples tested for PFOA, PFNA, PFDA, PFUnDA, PFHxS, PFHpS, and PFOS, all detected at LOD = 10 ng/L with PFOS, PFOA, and PFHxS being found in > 70% of the samples (24). In line with our finding of high PFOS and PFOA detection frequency, previous studies have consistently reported a widespread detection of these congeners in human milk (10, 16, 23, 24). We also found that PFHxS and PFHxA were detected in 71.6% and 70.0% of the samples, respectively; yet their detection frequencies in human milk have been inconsistently reported in U.S. studies. Zheng et al. found detection frequencies of 90% for PFHxS and 64% for PFHxA in 2019 samples from women in Seattle, Washington, while Criswell reported that both chemicals were found in less than 8% of samples collected from 2014 to 2019 in New Hampshire women (10, 16). Similar variation was also found in Asian, European, and Canadian studies (25–27).

### Human Milk PFAS Concentrations

We found that PFOA had the highest concentration in human milk, followed by PFOS, PFHxA, PFHxS, and ADONA. In NHBCS, Criswell et al. reported PFOA levels comparable to our estimates (median: 17 ng/L) but found higher PFOS concentrations (median: 24 ng/L) (16).

In Seattle, Washington, Zheng et al. observed lower PFOA levels, but reported higher PFOS concentrations (10). With the exception of two recent studies from China and Spain, others have reported higher human milk PFOS levels than IMPRINT at earlier time points (10, 16, 26–29). PFHxA, the third most abundant PFAS in our study, is a prominent short-chain alternative to long-chain PFCAs, such as PFOS and PFOA (10). It is a PFAS that has been used in carpets, food packaging or other household products (30), In animal and *in-vitro* models, PFHxA has been shown to induce oxidative stress, endocrine disruption, liver and immune toxicity, while epidemiologic studies on PFHxA associated health effects in humans are lacking (31). PFHxA levels reported in our study (median: 10.44 ng/L) were comparable or higher than in other studies with human milk sampled about the same time (median: 9.69 ng/L in Seattle, WA and 1.58 ng/L in Spain); in China, South Africa, and Canada studies, PFHxA was detected in <50% of human milk samples (10, 23, 25, 26, 29, 32). PFHxA concentrations higher than in IMPRINT were found in two Korean studies (medians: 47 ng/L and 33 ng/L) (33, 34). Among the PFAS with the highest human milk concentrations in our analysis, PFHxS has been reported to bioaccumulate to a greater extent than PFOS and PFOA, and has been associated with immunotoxicity, neurotoxicity, liver toxicity, and endocrine disruption in human and animal studies (35, 36). Human milk PFHxS levels in IMPRINT were lower than in other U.S. studies (10, 16) .

### PFECA and Other Novel Alternatives to Legacy PFAS in Human Milk

Our study was the first to measure PFECA in U.S. human milk. We observed that ADONA was detected in 68% of the samples (median: 3.5 ng/L), whereas the other PFECAs were detected in < 5% of samples. The only previous human milk study that detected ADONA was from samples collected in China and Sweden, which reported a 30% detection frequency in the samples from the Shanghai and Jiaxing provinces collected in 2015-2016 but found that it was undetected in most samples collected in Stockholm in 2016 (37). In milk samples where ADONA was detected, the study reported levels comparable to those in IMPRINT (3 ng/L versus 3.5 ng/L) (37). ADONA is a 3M polymer processing aid (PPA) and has been used as a PFOA alternative since 2008 for it has been suggested to bioaccumulate less than PFOS or PFOA but its potential toxicity in humans is unknown (37). Yet, in male rabbits, ADONA administered at 0.1 mL for eye and 0.5 mL for skin of 30% stock solution caused moderate to severe eye irritation and mild skin irritation (38). In male rats, ADONA was a possible PPARα agonist at 30 mg/kg administered at single oral and intravenous doses, and the chemical’s primary target was the liver in males and the kidney in females (38). Our study included other novel PFAS alternative chemicals such as HFPO-DA (or GenX), 9Cl-PF3ONS also known as 6:2 Cl-PFESA or the commercial name ‘F53B’, and 11Cl-PF3OUdS (or 8:2 Cl-PFESA), all of which were detected in ≤ 1% of samples.

### Human Milk PFAS Predictors

Only two previous studies examined human milk predictors of PFAS concentrations (18, 39) One included 35 samples collected from women in Lebanon between 2018 and 2021 and identified dairy products, egg consumption and tap water use for drinking as predictors of higher human milk total PFAS (39). The other was the NHBCS, which reported that low parity and the duration of previous lactation predicted higher milk PFOA and PFOS concentrations; fewer postpartum weeks at milk collection predicted higher milk PFOA concentrations, while low pre-pregnancy BMI predicted higher PFOS (16). Consistent with our results of high socio-economic status being associated with high PFHxS and ADONA, previous studies reported high socioeconomic status and being non-Hispanic White as predictors of high serum levels of legacy PFAS (40, 41). Fabelova et al. reported serum PFAS levels in 5,897 pregnant women from nine European cohorts and found that older maternal age was a predictor of higher levels of PFOA, PFOS, PFHxS and PFNA in maternal and cord blood after adjusting for parity, smoking, previous breastfeeding, and food consumption (42). In line with our findings, multiple pregnancies, menstruation, and breastfeeding have been suggested to be major PFAS elimination routes from mothers, with the chemicals being transferred from the mother to the child or via placental blood loss at childbirth (42). The average PFOS and PFOS placental transfer from the mother to the fetus has been estimated to be 13.7 and 8.32 ng/day respectively, resulting in a daily reduction in the PFAS body burden through pregnancy of 30.1 and 11.4 ng/day, respectively (43). We found sampling season to be a strong predictor of human milk PFAS, and the reason for this is unclear. Consistent with potentially higher exposure to PFOA, PFHxA and ADONA from water sources in the Fall compared to Winter or Spring, seasonal variations in PFAS exposure from source water have been reported (44). In cold regions, it has been suggested that groundwater is additionally contaminated with PFAS from soil mobilized by snow melt, increasing PFAS exposure through water sources at seasons with elevated temperatures (44). Moreover, higher temperatures enable PFAS volatilization and may contribute to increased exposure (45).

### Limitations and Strengths

Our study had limitations. The sample size of 100 participants is relatively small, and mothers recruited from the greater Cincinnati Metropolitan area may not be generalizable to the U.S. or other parts of the country. Our analysis of human milk PFAS predictors did not include important variables such as the main drinking water source and diet or consumption of contaminated food. PFAS were measured in human milk sampled at a single time point; there is evidence that human milk PFAS concentrations may vary over the course of lactation and breastfeeding. A Swedish study reported a significant PFOA decrease over the course of lactation, whereas PFHxS and PFOS levels did not vary (24). PFAS with a higher transfer efficiency may be associated with reduced maternal PFAS burden that can lead to faster decline in PFAS in human milk (24). Also, this analysis of PFAS in human milk used targeted analyses of PFAS with known standards, whereas using non-targeted analyses could lead to examination of new or unexpected PFAS. Non-targeted analyses can allow for enhanced understanding of exposure by allowing for the discovery of new congeners that are not measured with targeted analyses. Nonetheless, our study had strengths. It is one of the few reports on human milk PFAS from U.S. participants. Also, it is one of few studies where milk was collected at a standardized time (6 weeks), enabling comparison of risk factors between mothers. It is also the first study to determine PFAS EDI to infants from human milk and to determine TWI. The milk samples were analyzed for 29 different PFAS and included legacy as well as emerging PFAS that have not been previously adequately investigated. Our sample also consisted of a racially diverse population, while the past U.S. studies included only non-Hispanic White participants (10, 16, 17).

## Conclusions

Human milk samples collected between 2019 and 2020 in the Greater Cincinnati, Ohio, area from nursing women who participated in the IMPRINT study contain several legacy and emerging PFAS and transferred to children at levels above tolerable limits (TWI) established by the EFSA. Future studies with non-targeted PFAS measurements could inform the public of exposures from yet to be known PFAS exposures. Future studies could also include repeated measures of human milk PFAS over the course of lactation. Future comparisons of milk with serum from the same women at the same collection time are warranted to investigate a partitioning coefficient that could inform the public of the potential health effects of these chemicals in breastfed children.

## Data Availability

All data produced in the present work are contained in the manuscript and the online supplement

## Acknowledgement

We would like to thank all the participants in the IMPRINT study for making this work possible.

